# Changes in non-linear and time-domain heart rate variability indices between critically ill COVID-19 and all-cause sepsis patients – a retrospective study

**DOI:** 10.1101/2020.06.05.20123752

**Authors:** Rishikesan Kamaleswaran, Ofer Sadan, Prem Kandiah, Qiao Li, James M. Blum, Craig M. Coopersmith, Timothy G. Buchman

## Abstract

**Objective:** To measure heart rate variability metrics in critically ill COVID-19 patients with comparison to all-cause critically ill sepsis patients.

**Design and patients:** Retrospective analysis of COVID-19 patients admitted to an ICU for at least 24h at any of Emory Healthcare ICUs between March and April 2020. The comparison group was a cohort of all-cause sepsis patients prior to COVID-19 pandemic.

**Interventions:** none.

**Measurements:** Continuous waveforms were captured from the patient monitor. The EKG was then analyzed for each patient over a 300 second (s) observational window, that was shifted by 30s in each iteration from admission till discharge. A total of 23 HRV metrics were extracted in each iteration. We use the Kruskal–Wallis and Steel–Dwass tests (p < 0.05) for statistical analysis and interpretations of HRV multiple measures.

**Results:** A total of 141 critically-ill COVID-19 patients met inclusion criteria, who were compared to 208 patients with all-cause sepsis. Demographic parameters were similar apart from a high proportion of African-Americans in the COVID-19 cohort. Three non-linear markers, including SD1:SD2, sample entropy, approximate entropy and four linear features mode of Beat-to-Beat interval (NN), Acceleration Capacity (AC), Deceleration Capacity (DC), and pNN50, were statistical significance between more than one binary combinations of the sub-groups (comparing survivors and non-survivors in both the COVID-19 and sepsis cohorts). The three nonlinear features and AC, DC, and NN (mode) were statistically significant across all four combinations. Temporal analysis of the main markers showed low variability across the 5 days of analysis, compared with sepsis patients.

**Conclusions:** Heart rate variability is broadly implicated across patients infected with SARSCoV-2, and admitted to the ICU for critical illness. Comparing these metrics to patients with all-cause sepsis suggests a unique set of expressions that differentiate this viral phenotype. This finding could be investigated further as a potential biomarker to predict poor outcome in this patient population, and could also be a starting point to measure potential autonomic dysfunction in COVID-19.

## Introduction

The novel coronavirus (SARS-CoV-2) has led to a large cascade of transmissions resulting in high numbers of individuals hospitalized for the coronavirus disease 2019 (COVID-19), an impact still being accounted across the globe. A major target of the disease has been the respiratory components, resulting in acute respiratory insufficiency and failure. In patients who require mechanical ventilation, reported mortality rates exceed 50% (1, 2). However, COVID-19 is not only a respiratory disease. Cardiac, renal, hemodynamic, hematological and neurological manifestations were noted in critically-ill COVID-19 patients (3–5). The multi-system injury and high mortality rate requires means to identify patients at high risk.

Heart rate variability (HRV) is a naturally occurring phenomena, which takes different patterns in critical illness (6). HRV has long been associated to be a surrogate measure of cardiac autonomic tone (7–10). Analyzing complex dynamic features from the electrocardiogram (EKG), has been shown to indicate early cardiorespiratory complications (11, 12), autonomic dysfunction (13), sepsis (14) and death (15). These HRV measures, drawn from both temporal and frequency domain, have also been used in predictive models for early and rapid identification of deterioration in the ICU (16–20). However, the exploration of a comprehensive list of HRV metrics have not been investigated among COVID-19 patients. Therefore, in this study we explore the association and implications of these components in critically ill COVID-19 patients, identify features that differentiate survivors and non-survivors. Separately, we study the similarities of these characteristics to non-COVID-19 all-cause sepsis patients in a multi-ICU, single healthcare system.

## Methods

The study was approved by the Emory University Institutional Review Board. All medical, surgical. neurocritical, transplant and cardiac ICU admissions with COVID-19 between March 1 through April 31^st^, 2020 within Emory Healthcare system were screened. Patients were selected to be included in the analysis if they were in the ICU for greater than 24 hours. Controls were identified as all-cause sepsis patients meeting Sepsis-3 criteria (21), between 2015-2017, a previously published patient series (20). Patients with less than 5 days of ICU admission were excluded. Continuous bedside monitoring data was extracted during the patient’s ICU stay, and heart rate variability measures were generated using a sliding window. The data was exported from the archival system, de-identified and analyzed using open-source and proprietary statistical programs.

## Data Abstraction

Continuous waveforms were captured from the General Electric (GE) bed side monitors across Emory affiliated hospitals, using the BedMaster system (Excel Medical Electronics, Jupiter FL, USA). The data archival infrastructure is available in 152 beds spanning medical, surgical, transplant and cardiac ICUs across the health system. For this study, continuous EKG were captured from the bedside, each sampled at a sampling frequency of 250 Hertz (Hz). The EKG was then analyzed for each patient over a 300 second (s) observational window, that was shifted by 30s in each iteration from admission till discharge. A total of 23 HRV metrics were extracted in each iteration using the ‘PhysioNet Cardiovascular Signal Toolbox’ (22). The list of measures used in for this study is numbered in Table 3. Where any 300s observational window had more than 20% of poor data quality, the segment was discarded from the analysis.

## Statistical Analysis

MATLAB® was used to analyze the continuous EKG and extract relevant HRV features. The statistical analysis of this data was analyzed using Python Scikit-learn (23) and JMP ® (SAS institute) software. Demographics were evaluated for statistical significance (p < 0.05) using the chi-square method for categorical and standard Student’s t-test or one-way analysis of variance (ANOVA) for continuous variables. We use the Kruskal–Wallis and Steel–Dwass tests (P <0.05) for statistical analysis and interpretations of HRV multiple measures.

## Encounter Level Analysis

Due to the use of a 300s sliding window, the average number of observations sampled per patient were in excess of 10,500. In order to estimate encounter level variability, we derived the average value for each HRV measure. Since the goal of the study is to detect early marker of deterioration, we limit the period of analysis to the first five days (120 hrs.) from ICU admission. Histograms were generated using Scikit-learn for each measure, and separated by the class label.

## Trajectories of Disease

Temporal trajectories of each patient were generated to investigate differences in acuity and deterioration magnitude among COVID-19 survivors and non-survivors. Aggregate features of HRV were generated by uniformly sampling data across each 8-hour segment from admission until the fifth day. In-hospital mortality outcome was retrieved from the clinical record.

## Results

A total of 778 hospitalized patients were identified in a clinical chart review for positive SARSCoV-2 tests by use of quantitative RT-PCR (qRT-PCR). From this cohort, 413 encounters were mapped to bedside monitors that were actively archiving data during the time of their hospitalization. We excluded 272 patients due to insufficient data or poor quality (>20% missing data) data from the first five days of ICU admission, leaving a total of 141 patients who were analyzed. We identified 557 encounters, pre-COVID-19, who met sepsis-3 criteria during their ICU stay and had high-frequency bedside monitoring captured. 208 of those encounters had sufficient data quality in the first five days of ICU stay, and eligible for the study. Table 1 presents a description of the clinical and demographic characteristics of the cohort. Notably, the proportion of African American patients in the COVID-19 cohort was over twice as the proportion in the sepsis cohort. Within both groups, the non-survivors were older on average, compared to the survivors.

A basic description of the different HRV indices of the COVID-19 and Sepsis group is detailed as a distribution histogram plot in Supplemental Figure 1. While the total number of encounters in the sepsis group are greater, there was significantly more data available per encounter in the COVID-19 group. Prior to inclusion criteria being applied, there were approximately 680,799 seconds (∼8 days) of data points on average per encounter in the COVID-19 group, while 206,288 seconds (2.4 days) in the sepsis group.

Descriptive statistics of the HRV measures are detailed in Table 2. Notably, both Kruskal–Wallis and Steel–Dwass tests for multiple comparisons show statistically significant separations between the COVID-19 and all-cause sepsis groups. Specifically, on seven specific markers, which consists of three non-linear markers, including SD1:SD2, sample entropy (SampEn), approximate entropy (ApEn) and four linear features mode of Beat-to-Beat interval (NN), Acceleration Capacity (AC), Deceleration Capacity (DC), and pNN50, were statistical significance between more than one binary combinations of the sub-groups (comparing survivors and non-survivors in both the COVID-19 and sepsis cohorts). The three nonlinear features and AC, DC, and NN (mode) were statistically significant across all four combinations. A number of Beat-to-Beat interval metrics show significant QRS complex elongation between COVID-19 and sepsis, with the NN (mean) appearing on average, greater among both COVID-19 sub-groups than in sepsis survivors and non-survivors.

Figure 1A illustrates a box-plot of the six highly differentiating measures, with their respective distributions rendered by a violin plot, illustrating kernel density in the background. Figure 1B illustrates correlation matrix of each HRV measure value, correlations were observed among metrics relating to vagal tone (RMSSD, pNN50, HF, SD1). Non-linear components consisting of SD1:SD2, SampEn, and ApEn was the most distinguishing factor between each sub-group, demonstrates a bimodal distribution within the COVID-19 survivors and non-survivors, which was not observed among the all-cause sepsis cohort.

**Figure 1:**
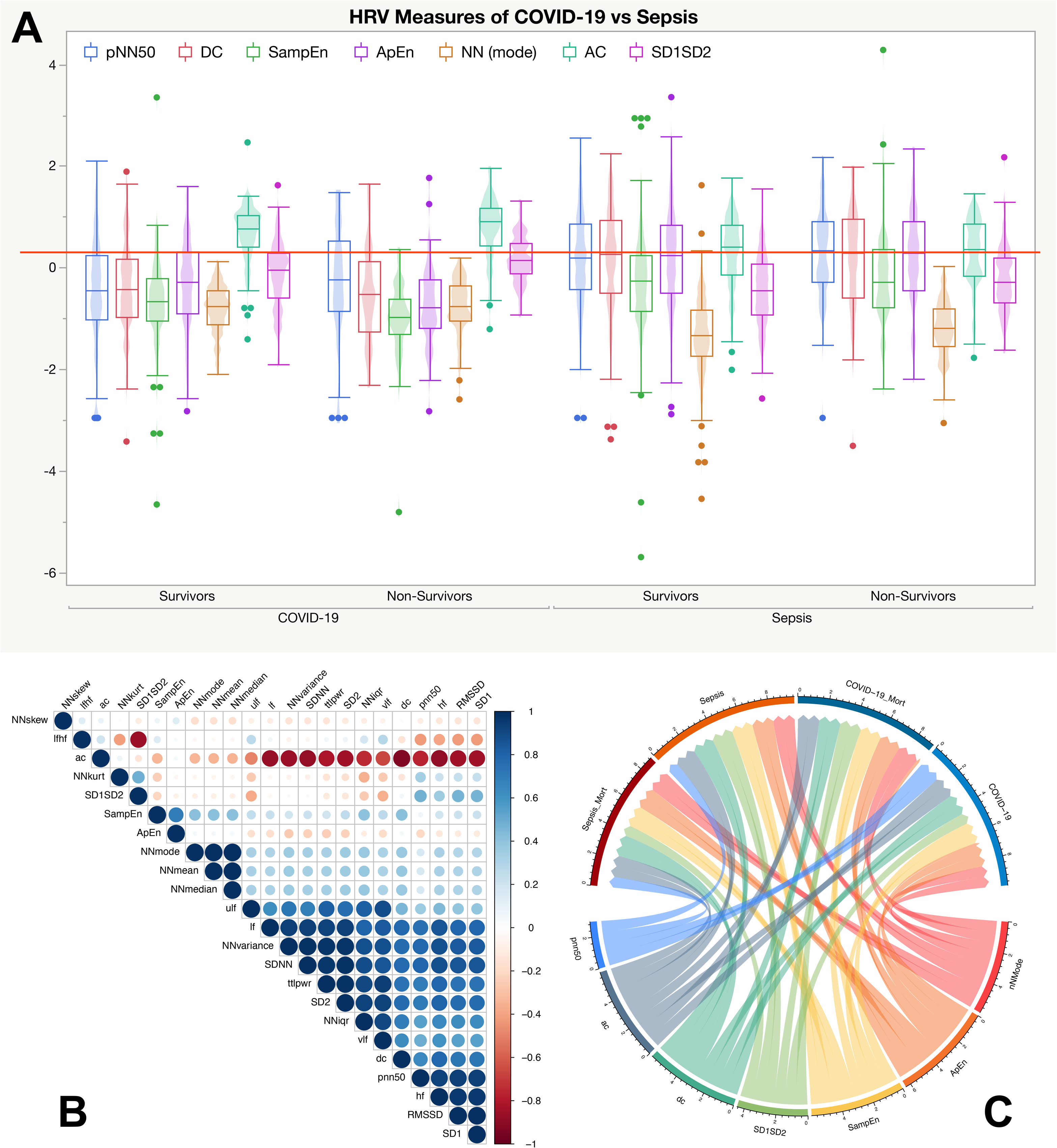
A. Illustrates box-plots with violin subcomponents characterizing the distribution within each group. **B**. A correlation plot of all HRV measures, suggesting strong interactions between SDNN, pNN50, LF, VLF, and SD1, previously linked to parasympathetic components.**C**. A chord diagram illustrating interactions among HRV measures and the four unique classes of COVID-19 and sepsis survivors and non-survivors. The upper portion of the chord diagram lists the four sub-groups, survivors and non-survivors of COVID-19 and sepsis. The lower portion illustrates the six HRV measures which demonstrated statistically significant separation between the groups. The size of the edge in the lower portion represents more statistically significant interactions across each group, along with the magnitude of the statistical significance, i.e. p< 0.001 are weighted higher.

AC and DC components were significantly different among the two broad groups, with COVID-19 groups having a lower DC but a greater AC median value compared to the sepsis groups. pNN50 in COVID 19 groups were on average lower in comparison to sepsis groups, where sepsis non-survivors had the largest pNN50 median. The correlation plot (Figure 1B) further shows high correlations among the beat-to-beat (NN*) derived features and HRV components attributed to parasympathetic activity. AC shows negative correlation with many of the time-frequency and nonlinear features, however no collinearity were found in ratios thought to represent sympathovagal balance (SD1:SD2, LF:HF, SampEn, ApEn). LF:HF ratio were found to have elevated median values between COVID-19 and all-cause sepsis, however is not distinguished when considering readings in non-survivors.

Figure 1C illustrates a chord diagram of the interactions among the HRV measures with significant statistical significance across COVID-19 and all-cause sepsis cohorts using the Kruskal–Wallis and Steel–Dwass tests (P < 0.01). The three non-linear measures, along with AC DC, and NN (mode) had the greatest number of interactions (width of the link) among each of the four groups with a p value < 0.001. pNN50 had fewer strongly significant interactions, and could not differentiate COVID-19 non-survivors from survivors. SD1:SD2 had stronger statistical significance among Sepsis survivors and COVID-19 non-survivors.

Temporal trajectories of values were accumulated over each 8-hr segment of data and projected from admission until day 5. Figure 2 illustrates the normalized temporal trajectories for the six most statistically significant HRV measures between the cohorts, the 95% confidence interval (CI) is marked by the shaded region around each line. The mode of NN, pNN50, ApEn, and SampEn starts lower in the COVID-19 cohort and after 5 days remains lower than the sepsis cohort. While DC and SD1:SD2 begin higher in the COVID-19 cohort and end lower during the same timeframe. AC, DC, pNN50, NN (mode), the 95% CI is much tightly bound in COVID-19 patients. While among the non-linear metrics the 95% CI bound is much broader. Within all seven HRV measures, the sepsis cohort demonstrates dynamic fluctuations over their ICU stay that are not observed within the COVID-19 patients.

**Figure 2:**
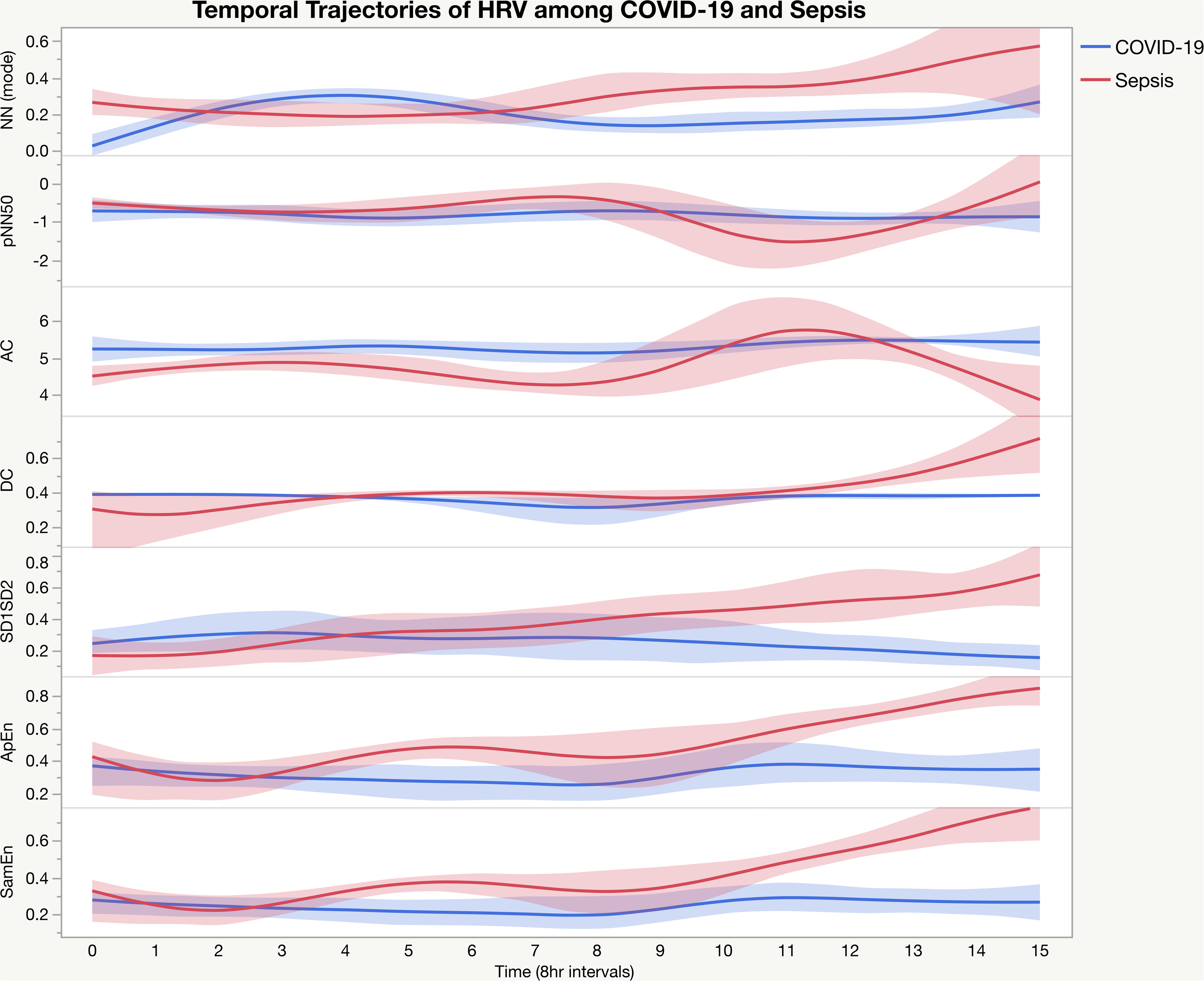
Temporal trajectories among highly significant features distinguishing COVID-19 and sepsis cohorts. The 95% confidence interval is illustrated as shades bounding each line. Unlike in the sepsis cohort, where a dynamic change is observed in these HRV measures, the COVID-19 group has more monotonic characterization.

## Discussions

By comparing retrospective cohorts of COVID-19 and non-COVID-19 all-cause sepsis patients, this study demonstrates a distinctively expressed subsets of nonlinear and time domain HRV measures. The results show marked separation between sepsis and COVID-19 patients, and also separates between survivors and non-survivors.

When comparing the cohorts from a demographic stand point, there were significantly more African Americans in the COVID-19 dataset, which is consistent with recent reports (24). There was no meaningful difference between mortality among ethnicities. No distinctive differences by sex was observed in outcomes, with about an equal number of male non-survivors (52%).

Out of the battery of HRV indices analyzed, only some were able to separate the groups (COVID-19 and sepsis) and subgroups (survivors and non-survivors) in a statistically significant manner. The most statistically significant measure of HRV across multiple-groups was ApEn, SampEn and SD1:SD2. ApEn derived from methods of information theory, is a measure of the degree of nonstationarity, and higher values indicate poor irregularity and thus suggested a poor parasympathetic tone. SampEn, is a similar measure of signal complexity as ApEn, however a more robust one, when considering shorter observational time. Both SampEn and ApEn were lower in the COVID-19 cohort than in all-cause sepsis, indicating a reduced dynamic in heart rate modulation.

Of the significant *time-domain* HRV indices we measured (Table 3), pNN50, AC, DC, and NN (mode) strongly distinguished COVID-19 patients from all-cause sepsis. Cardiovascular complications within COVID-19 has been documented in many recent works (25–27). In our results we see on average a greater NN value (lower heart rates) within COVID-19 patients when compared to all-cause sepsis. Apart from NN (mode), most beat-to-beat metrics show a higher baseline distance among COVID-19 patients. IQR of NN shows that COVID-19 profiles among survivors and non-survivors are closer to all-cause sepsis non-survivors, indicating a potential increase in risk for mortality.

pNN50 has long been a predictor of poor cardiac physiology (28). In our analysis, we see that after day 2 (7^th^ block, pNN50, Figure 2), the mean pNN50 distinguishes from earlier periods and has a distinct difference progressively during the ICU stay. COVID-19 seems to significantly decreased DC, while AC was higher (Figure 1). Increased AC and decreased DC has specifically been shown to be predictive of mortality in myocardial infraction (29) and heart failure (30), and both AC and DC components were implicated in inflammatory mediation (31) and indicate significant vagal activation (32). In particular, decreased DC has been shown to be linked with impaired PNS activity (33). Interestingly, evaluating temporal trajectories also emphasize distinct characteristics between the groups, while the trajectory of values among the COVID-19 group did not change significant over time, in contrast a greater degree of temporal dynamics was observed in the sepsis cohort (Figure 2).

The ratio of SD1:SD2 derived from the Poincaré Plot, a visual geometric measure of self-similarity in periodic functions. SD1 has been correlated with the baroreflex sensitivity and measures short-term changes modulated by respiratory sinus arrhythmia (RSA) associated to parasympathetic activity (34). The SD1:SD2 ratio is a measure of autonomic balance, whereby decreased SD1:SD2 ratio indicates an elevated sympathetic tone and suppressed parasympathetic activity (35). It was interesting to note that in our dataset the COVID-19 group had a marginally higher SD1:SD2 ratio than all-cause sepsis, in contrast to findings from DC, pNN50, SampEn and ApEn which indicate a lower parasympathetic tone. An elevated SD1:SD2, may indicate a broad and complex dynamics between the sympathetic and parasympathetic arms, and/or medication influence being reflected in COVID-19.

There are several pathophysiological explanations for the differences shown between COVID-19 and sepsis patients. One option relates to a direct cardiac injury. Indeed, cardiac manifestations in COVID-19 patients are common (5). A direct injury could theoretically damage the cardiac pacer or the conduction system, resulting in changes of HRV. A second option to be considered is an injury to the autonomic system. Many of the parameters found to differentiate the groups in this cohort, were previously correlated with changes in the sympathovagal tone (12, 13). The level of neuro-tropism of SARS-CoV-2 remains unclear, and specifically whether or not it could invade parasympathetic fibers via the gastrointestinal tract or the lungs.

A third option could be related to the binding of SARS-CoV-2 to ACE2, resulting in the loss of the protective pathway against a dysregulated autonomic system (36). The loss of ACE2 function has been associated to binding of the SARS-CoV-2 virus driven by endocytosis and initiation of the proteolytic cleavage and processing. ACE2 regulates the RAS system by converting Ang I and Ang II into Ang 1–9 and Ang 1–7, respectively (37, 38). Loss of function of the ACE2 receptor, and thus an unregulated Ang II has been associated with hypertension and cardiovascular autonomic dysfunction (39). In a cohort of 12 COVID-19 patients, circulating Angiotensin II was found to be significantly elevated when compared to controls, and showed linear relationship with viral load (40). Ang II exerts several actions on the sympathetic arm of the autonomic nervous system, facilitating increased sympathetic outflow and neurotransmission.It also acts on the baroreflex mechanism to modulate blood pressure (41, 42), and therefore by surrogate influence the cardiac non-stationarity. Finally, the targeting of the respiratory system by the SARS-CoV-2 virus impacts breathing rate, and thus the respiratory sinus arrhythmia (RSA), which in turn modulates high-frequency activities of the cardiac system.

The combination of the seven non-linear and temporal HRV measures indicates potential sympathovagal imbalance within this pilot study, with various degrees of interaction between the sympathetic and parasympathetic arms. Among COVID-19 patients, there were more pathological regularity observed both overall and temporally. This indicates potential suppressed parasympathetic activity that may be driven by interference with the RAS system. Parallel influences from respiratory components, affecting the RSA further contribute to a dysfunctional autonomic regulation of the cardiac system. These cues, as shown within this analysis, can be distinctive between the cohorts, including between survivors and non-survivors, and therefore potentially predictive of mortality.

## Limitations

There are a number of limitations of this study, including the fact that only 141 COVID-19 patients were eligible for inclusion. Furthermore, the analysis was performed on a cohort from multiple hospitals and units, yet still from a single health system in a single metropolitan. COVID-19 related mortality in our health system was lower than prior reports which could affect generalizability. This study was an observational cohort study performed retrospectively, and therefore causality analysis is limited. Specifically, this study did not examine the correlation between the HRV metrics and medications used to treat the patients. Additional work needs to be done to evaluate the performance of these measures prospectively. Developing methods to automatically capture this data and generate point-of-care biomarkers is an active area of ongoing work (43, 44).

## Conclusion

Heart rate variability is broadly implicated across patients infected with SARS-CoV-2, and admitted to the ICU for critical illness. These biomarkers may suggest a degree of autonomic dysfunction that can be longitudinally expressed. Key subsets of measures were identified across time, frequency and nonlinear domains. The results highlight potential biomarkers that could separate COVID-19 patients from all-cause sepsis patients. More importantly, these results prove preliminary data to allow early separation of survivors and non-survivors. Temporal trajectories of these markers further suggest significant decoupling as the disease progresses, with salient decoupling noticeable as early as at ICU admission. While the results of this study are early, we establish the premise that these higher-dimensional features of heart rate can be associated with poor outcomes among COVID-19 patients.

## Data Availability

Data is not available for public release, secondary data abstractions can be requested to the corresponding author.

## Acknowledgements

This work was partially funded by the Surgical Critical Care Initiative (SC2i).

## List of Figures

**Supplemental Figure 1:**
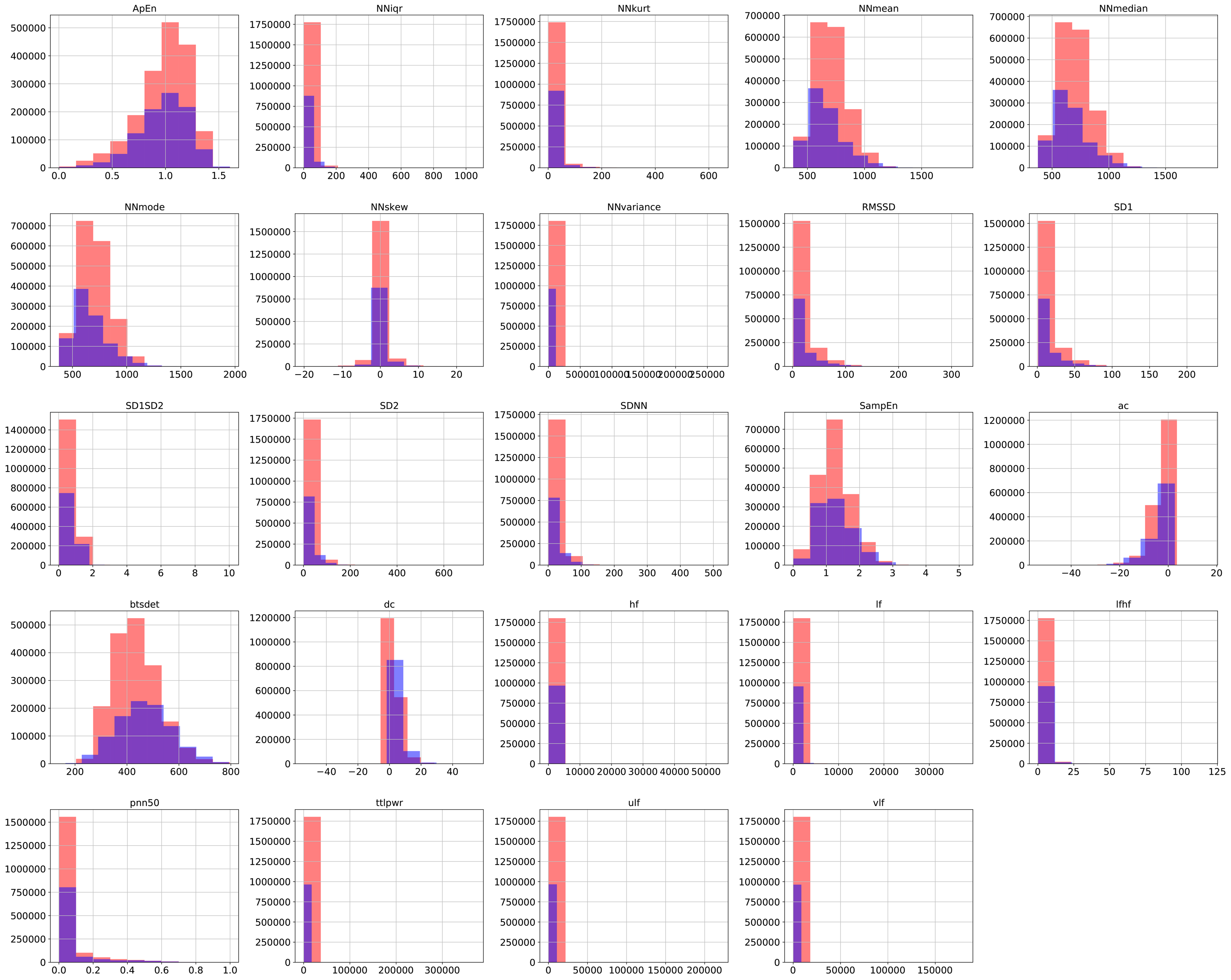
A stacked histogram of HRV measures calculated across COVID-19 patients (orange) and Sepsis (blue).

